# Bias Reduction through Analysis of Competing Events (BRACE): A Novel Method to Mitigate Bias from Residual Confounding in Observational Data

**DOI:** 10.1101/2020.12.18.20248507

**Authors:** Loren K. Mell, Tyler Nelson, Caroline A. Thompson, Casey W. Williamson, Lucas K. Vitzthum, Jingjing Zou

**Affiliations:** Department of Radiation Medicine and Applied Sciences, University of California San Diego, La Jolla, California; La Jolla Center for Precision Radiation Medicine, La Jolla, California; Division of Epidemiology and Biostatistics, San Diego State University School of Public Health, San Diego, California; Division of Epidemiology, University of California San Diego Herbert Wertheim School of Public Health and Human Longevity Science, La Jolla, CA; Department of Radiation Oncology, Stanford University, Stanford, California; Department of Family Medicine and Public Health and Department of Mathematics, University of California San Diego, La Jolla, California

## Abstract

**Purpose:** To introduce a method to mitigate bias from residual confounding in non-randomized data and examine its performance under varying conditions using simulated data.

**Methods:** We developed a method called Bias Reduction through Analysis of Competing Events (BRACE) based on a proportional relative hazards model. We followed recommended guidelines (ADEMP) established for the conduct of simulation studies. The primary estimand of interest was the treatment effect on the composite hazard for a primary or competing event. We compared the BRACE method to a standard Cox proportional hazards regression model in the presence of an unmeasured confounder, using a parametric (Weibull) simulation model. We examined estimator distributions, bias, mean squared error (MSE), and coverage probability for both methods using ridge, box-and-whisker, forest, and zip plots, respectively. Comparisons with a hypothetical validation estimate treating the confounder as measurable were also performed.

**Results:** We presented 16 simulation scenarios under varying parameters. In simulations where residual confounding was present, the BRACE method uniformly reduced both bias and MSE compared to standard Cox models. In the scenario of moderate bias with an effective but non-toxic treatment, MSE was 3.51×10^−2^ with the standard model vs. 0.259×10^−2^ with the BRACE method. In the absence of bias, the BRACE method introduced bias toward the null (2.90 x10^−2^) compared to the standard method (0.331×10^−2^), albeit with lower MSE (0.341 x10^−2^ vs. 0.484 x10^−2^, respectively). Relative to the standard approach, the BRACE method markedly improved coverage probability, but with a tendency toward overcorrection in the case of the effective but non-toxic treatment. Conclusions were similar under different parameter assumptions.

**Conclusion:** The BRACE method can reduce bias and MSE in the setting of residual confounding.

## Introduction

Bias due to residual confounding (often called treatment selection bias) is an important consideration when drawing inferences from non-randomized comparative effectiveness studies.^1,2^ In the absence of randomization, multivariable regression models and propensity score models are commonly used approaches to obtain treatment effect estimates, by adjusting for effects of measured confounders.^3^ However, residual confounding from unknown and unmeasured confounders is still a pernicious problem that can undermine conclusions from such analyses and cannot be overcome by scoring and weighting methods.^4-8^

Competing event analysis is an underutilized method that allows for the identification of residual confounding problems in non-randomized data sources, particularly when the effect of a treatment on competing events can be bounded *a priori*. ^9^ For example, while the simple addition of a novel cancer treatment to a standard regimen may have no effect on or even increase mortality from non-cancer health events, such as cardiac disease, it typically would not *reduce* the incidence of such events. Despite this, in non-randomized data, competing event analysis may reveal a *lower* incidence of competing health events in the group receiving more intensive treatment, due to unmeasured confounding by more favorable underlying health characteristics in this group.^9^ This paradox can be observed even after appropriately controlling for measurable confounders, and can be a critical driver of observed effects on combined endpoints like overall survival (OS) or progression-free survival (PFS). This phenomenon, when present, can be regarded as diagnostic of residual confounding (to the extent the underlying assumption is valid).

While diagnosing the problem of residual confounding is useful, this still leaves investigators with the question of what to do about it. In this study, we propose a method to mitigate bias due to residual confounding, leading to improved identification of the treatment effect when the method assumptions are valid.

## Methods

### Proportional Relative Hazards Model and Proposed Correction Model

Let Θ be the effect of a binary on the time-dependent (t) baseline hazard for a combined endpoint (e.g., progression-free survival) (*λ*_0_(t)). We can express *λ*_0_(t) as a function of the cause-specific hazards for two mutually exclusive events: event 1 (*λ*_01_(t)) the primary event (or set of primary events) and event 2 (*λ*_02_(t)), the competing event (or set of competing events), i.e.:

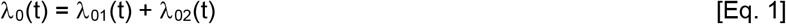

Dropping the argument t for simplicity, we derive the following proportional relative hazards model, where Θ_1_ and Θ_2_ are the effects of treatment on event 1 and event 2, respectively, and *λ*_1_ and *λ*_2_ are the resulting hazards for event 1 and event 2 under treatment:

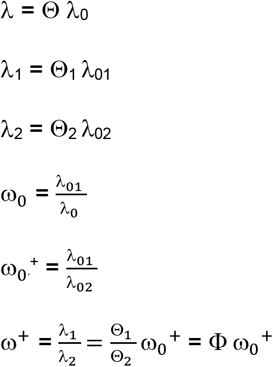

Where ω_0_^+^ and ω^+^ as the relative hazards for event 1 vs. event 2 under baseline and treatment conditions, respectively, and Φ as the effect of treatment on the ω_0_^+^ function. Θ, Θ_1_, Θ_2_, ω_0_, ω_0_P^+^ and ω^+^ are all non-negative hazard ratios. Here we assume ω_0_ is approximately invariant to time. Following Mell & Jeong,^10^ we get the following expression:

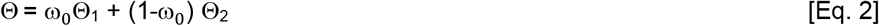

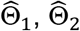, and 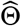, are the estimated treatment effects on event 1, event 2, and the combined event, respectively, and 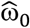 as the estimate of ω_0_. For the application of this method, we assume that Θ_2_ ≥ 1 (i.e., the treatment does not reduce the hazard for the competing event), treatment effects are independent of the baseline event hazards, and that 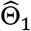 and 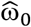 are unbiased 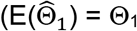and 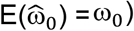. We bound the bias (ε) for Θ using the following derivation from [Eq. 1]:

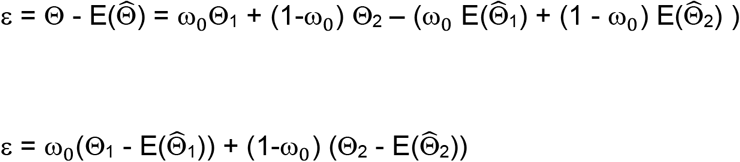

Therefore:

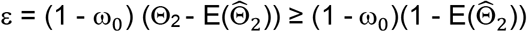

Thus we derive a partially corrected estimate of 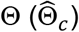 as follows:

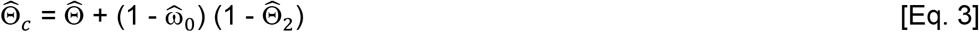

### Simulation Methods

Our methodology followed recommended guidelines (ADEMP) for the conduct of simulation studies.^11^ The primary aim of this simulation study was to compare estimates of a treatment effect on a combined endpoint (Θ) in the presence of treatment selection bias (i.e., ε > 0). We first employed a standard unadjusted Cox proportional hazards model^12^ to estimate the treatment effect 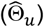, assuming the simulated confounding factor is unmeasurable. We then compared this estimate to the corrected estimate 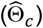 obtained using an alternative approach called Bias Reduction through Analysis of Competing Events (BRACE), based on the correction shown in [Eq. 3]. As a “validation” step, we also compared both methods to a hypothetical estimate obtained by re-weighting the Cox proportional hazard model with the inverse of a propensity score, treating the confounding factor as measurable.

For each method, we estimated and compared bias, mean squared error (MSE), and coverage probability using a parametric model under varying inputs for effect size, distribution parameter(s), sample size, and magnitude of bias. Effect sizes and bias were varied factorially, while the other model inputs were varied one-at-a-time. In varying effect size, we compared three conditions: no treatment effect (i.e., Θ_1_ = Θ_2_ = Θ = 1), effective but non-toxic treatment (i.e. Θ_1_ < 1, Θ_2_ = 1, Θ < 1), and effective but toxic treatment with negating effects on the composite endpoint (i.e. Θ_1_ < 1, Θ_2_ > 1, Θ = 1). The null hypothesis in all scenarios was Θ = 1 vs. the alternative hypothesis Θ ≠ 1. We applied the BRACE correction when 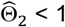 and the upper limit of the 95% confidence interval for 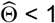; otherwise, 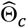 defaulted to 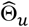. Note that the correction is applied to the estimate 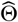, not 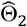.

For the parametrization of event times, we assumed a Weibull distribution for each of the cause-specific event times, with random censoring, under varying Weibull constants (*γ* < 1, *γ* = 1, *γ* > 1). We tested models using two sample sizes (large, N=1500 per trial; small, N=250 per trial). For the bias parameter, we tested three conditions: none, moderate, and severe). For moderate and severe bias conditions, the bias parameter was drawn from a Bernoulli distribution. The biasing factor increases the log odds of being assigned to the treatment group and decreases (by a constant multiple of 1/3) the baseline hazard for event 2 (*λ*_02_). Note that this factor does not directly influence the effects of treatment (because Θ_1_ and Θ_2_ do not vary with the factor), but does indirectly influence the composite treatment effect (Θ) due to its effect on *λ*_02_; for further discussion see Mell & Jeong.^10^

Simulated data were generated in SAS v9.4 (SAS Institute Inc, Cary, NC). For each scenario, we generated a sample of 500 trials. Random number seeds were set once per repetition and random number states were stored at the start of each repetition. For outcome regression modeling and bias corrections, we used R version 4.0.2, coxph function (*survival* package). For variance estimates of 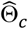, we used a Monte Carlo routine with 1000 replicates for the parameters 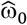 and 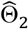. To estimate 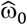 we used the Nelson-Aalen method, as described elsewhere.^13^ For graphical analysis of results, we used ridge, boxplot, zip, and forest plots (*rsimsum* and *ggplot2* package). All code is provided in the **Supplement**.

## Results

**Table 1** shows summary statistics (key comparisons highlighted) comparing treatment effect estimates with the standard Cox model vs. the BRACE method in the presence of unmeasured confounding. In the scenarios presented, the bias parameter (κ) increases the log odds of treatment in the presence of the confounder, while decreasing *λ*_02_ by a factor of 1/3. Thus, the confounder introduces heterogeneous risk for the baseline hazards for the competing and composite events (i.e., differing values of *λ*_01_ and *λ*_02_), without influencing the cause-specific treatment effects (Θ_1_ and Θ_2_).

**Table 1.**
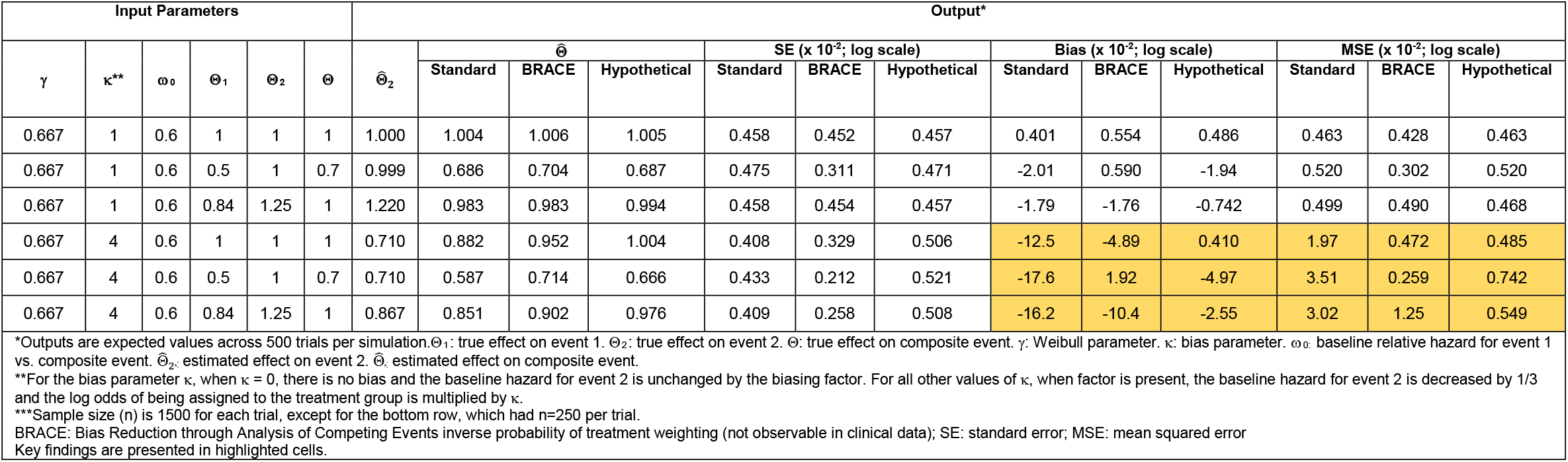
Summary statistics comparing standard Cox model vs. Bias Reduction through Analysis of Competing Events (BRACE) method in the presence of unmeasured confounding.

In general, the BRACE method uniformly reduced both bias and MSE compared to the standard approach. For example, in the case of moderate bias (κ = 4), with an effective but non-toxic treatment (**Table 1, row 5**), the MSE was 3.51×10^−2^ with the standard model vs. 0.259×10^−2^ with the BRACE method. In some cases, the BRACE method even reduced bias and MSE relative to the hypothetical, likely due its incorporation of prior knowledge about Θ_2_. For example, with an effective but non-toxic treatment (**Table 1, row 5**), the bias (−4.97×10^−2^) and MSE (0.742×10^−2^) were both greater with hypothetical case compared to the BRACE method.

Conclusions were similar under different assumptions about the Weibull parameter (*γ*), magnitude of bias, relative baseline event hazards (ω_0_), and sample size (except for standard errors (SEs) as expected in the small sample simulation) (**Supplementary Table**). In the scenario with no treatment selection bias and homogeneous risk for the baseline event hazards for each sample (**Supplementary Table, rows 1-3**), each method returns (as expected) unbiased estimates with low error. In the case of an effective but non-toxic treatment (**Supplementary Table, row 2)**, the BRACE method introduced bias toward the null (2.90 x10^−2^) compared to the standard method (0.331×10^−2^), albeit with lower MSE (0.341 x10^−2^ vs. 0.484 x10^−2^, respectively).

**Figure 1** compares ridge plots corresponding to simulations presented in rows 4-6 from **Table 1**. When the effect of treatment on the composite event was null (**Figure 1A** & **1C**), the distribution of the BRACE-adjusted estimator was intermediate between the standard estimator and the hypothetical. In the scenario with an effective but non-toxic treatment (**Figure 1B**), the BRACE method overcorrected slightly relative to the hypothetical, but with lower bias and MSE, as seen in **Figure 2** and **Figure 3**. In all cases, both bias and MSE were lower with the BRACE method compared to the standard estimator when residual confounding was present.

**Figure 1.**
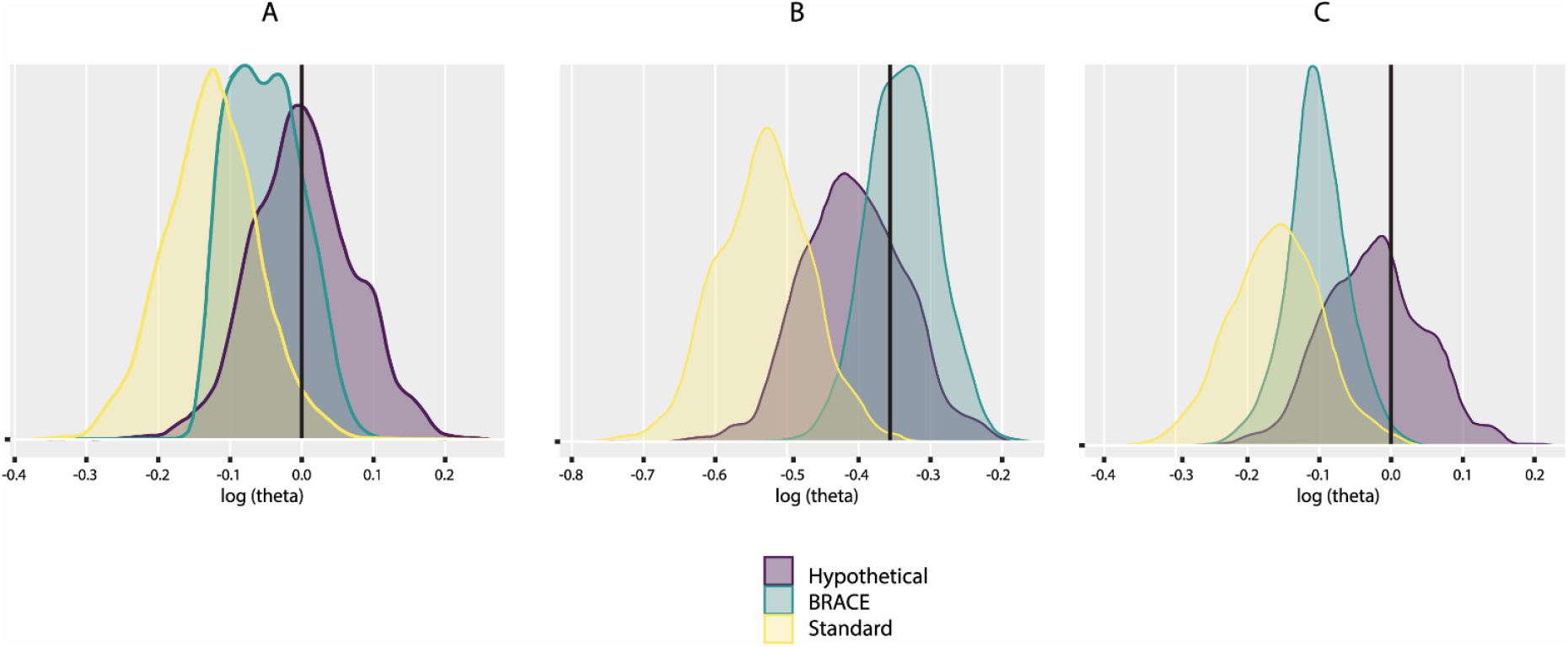
Ridge plots comparing standard vs. BRACE methods in the presence of residual confounding. The hypothetical case where the confounder is measurable is also shown for comparison. Figures A, B, and C correspond to rows 4, 5, and 6 from **Table 1**, respectively. (A) Null effects (B) Effective but non-toxic treatment (C) Effective and toxic treatment, with negating effects on the composite event.

**Figure 2.**
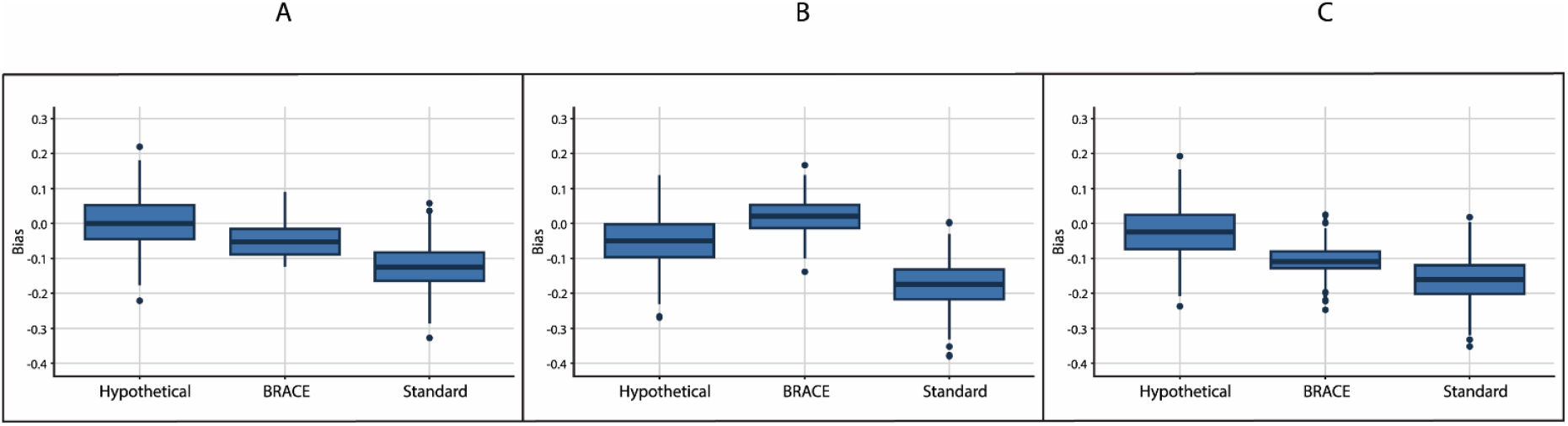
Box plots comparing bias (log scale) for the standard vs. BRACE methods in the presence of residual confounding. The hypothetical case where the confounder is measurable is also shown for comparison. Figures A, B, and C correspond to rows 4, 5, and 6 from **Table 1**, respectively. (A) Null effects (B) Effective but non-toxic treatment (C) Effective and toxic treatment with negating effects on the composite event.

**Figure 3.**
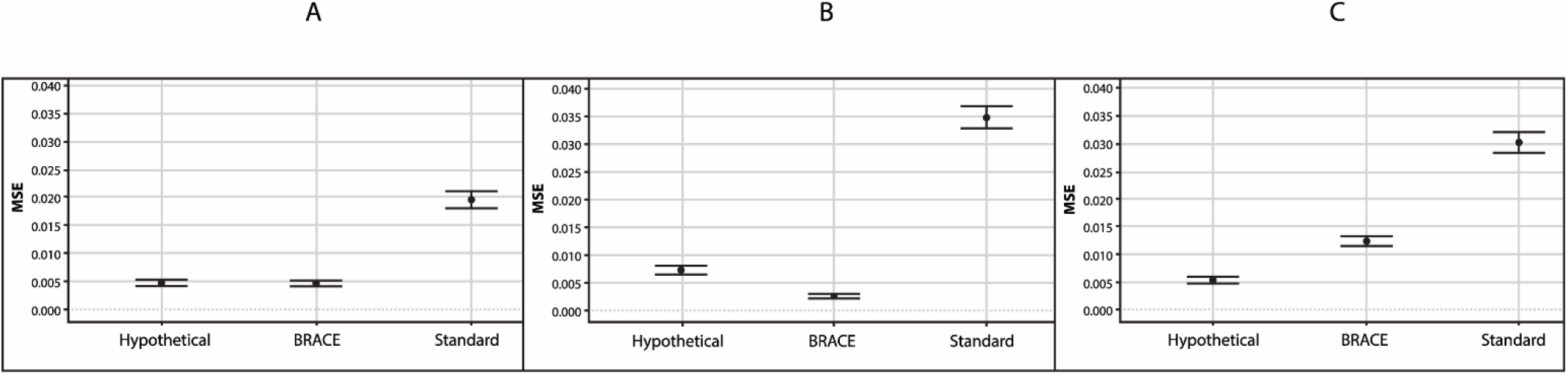
Forest plots comparing mean squared error (MSE) for the standard vs. BRACE methods in the presence of residual confounding. The hypothetical case where the confounder is measurable is also shown for comparison. Figures A, B, and C correspond to rows 4, 5, and 6 from **Table 1**, respectively. (A) Null effects (B) Effective but non-toxic treatment (C) Effective and toxic treatment with negating effects on the composite event.

**Figure 4** compares coverage probability for the standard, BRACE, and hypothetical approaches for the same set of simulations. Relative to the standard approach, the BRACE method markedly improved coverage probability (i.e., proportion of intervals containing the true value of Θ), but with a tendency toward overcorrection in the case of the effective but non-toxic treatment. For all scenarios, the BRACE method attenuated the tendency of the standard method toward extreme downward bias and improved inference regarding the likely true effect given the observed data.

**Figure 4.**
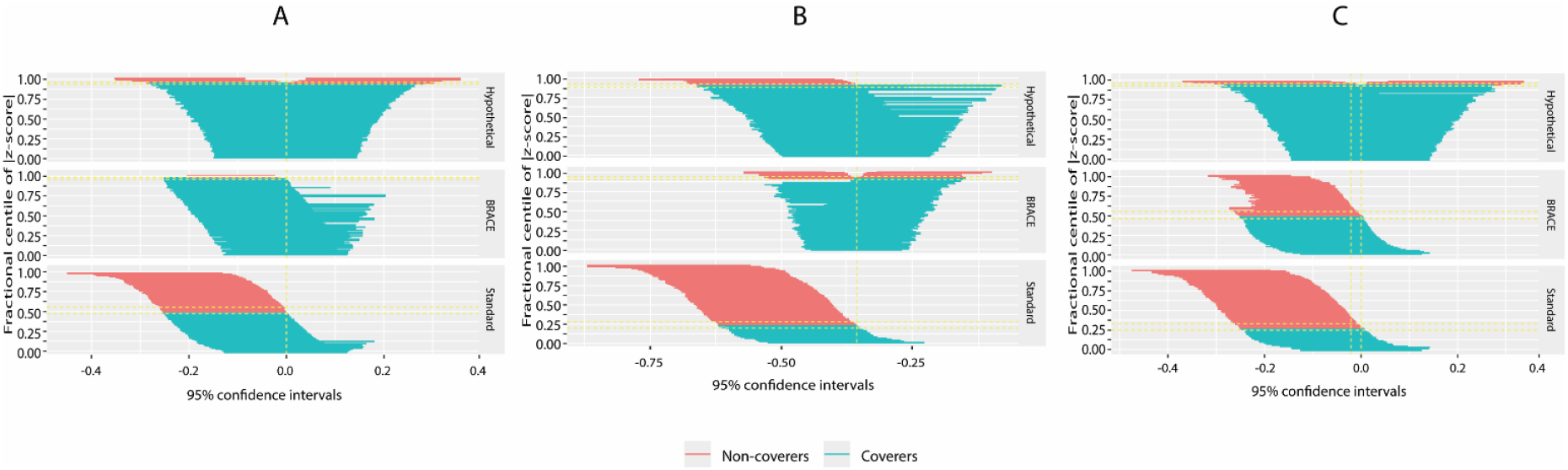
Zip plots comparing coverage probability for the standard vs. BRACE methods in the presence of residual confounding. The hypothetical case where the confounder is measurable is also shown for comparison. Figures A, B, and C correspond to rows 4, 5, and 6 from **Table 1**, respectively. (A) Null effects (B) Effective but non-toxic treatment (C) Effective and toxic treatment with negating effects on the composite event. Treatment effects are shown on the log scale.

In general, changing the input parameters had little effect on the ability of the BRACE method to improve estimation relative to the standard approach, as shown (to the extent practical) in **Supplementary Table**. In the primary analysis, we only applied the partial correction to trials for which 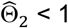 and the upper limit of the 95% confidence interval for 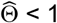. Another approach would be to apply the method to trials for which 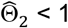 and 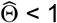, regardless of whether the null hypothesis is rejected. When we applied this less conservative approach using the same parameter inputs as in rows 4-6 from **Table 1**, we found that when all the treatment effects were null, expected values for bias (0.664×10^−2^) and MSE (0.219×10^−2^) were still low, with 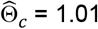. For the situations with an effective and non-toxic treatment and an effective but toxic treatment, the corresponding values were, respectively: bias: 1.92 x10^−2^ and −9.43×10^−2^; MSE: 0.26×10^−2^ and 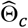: 0.714 and 0.911. All these expected values compared favorably to the standard approach. However, it should be noted that in the absence of treatment selection bias or effect heterogeneity, in the scenario with all null treatment effects, this strategy led to slight upward bias: 2.16 ×10^−2^; MSE: 0.318 ×10^−2^; 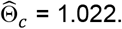.

## Discussion

Bias due to residual confounding (treatment selection bias) is a common problem affecting inferences from non-randomized observational studies in comparative effectiveness research. While multivariable regression and propensity score modeling are often used to mitigate bias, they cannot overcome the issue residual confounding, especially due to unobserved confounders. This problem frequently undermines the strength of conclusions from observational studies and can result in erroneous inferences.^1-9^ When effects of treatments on competing events can be bounded *a priori*, however, residual confounding can often be diagnosed using competing risks analysis.^9^

Building on a proportional relative hazards model, we derived a method to mitigate bias due to residual confounding when it is identified, resulting in lower model error compared to standard approaches. In some cases, due to the impact of effect heterogeneity, the BRACE method even reduced bias and MSE compared to a hypothetical model that could adjust fully for the confounding factor, likely due to the advantage of using prior information about treatment effects on the competing event. Nonetheless, it should be stressed that the relevant comparisons for clinical application are between the BRACE vs. standard method, since the hypothesized confounder is not directly measurable in real-world scenarios.

The gains using the BRACE approach depend on leveraging a critical, but plausible, assumption applicable to many comparative effectiveness studies in oncology: namely, that treatment does not reduce the hazard for competing health events. A typical example would be the addition of a new treatment to standard of a care (e.g. A vs. A+B design), where B denotes a potentially effective but potentially toxic treatment. With such designs, competing events may increase with the addition of B, either due to direct toxic effects, or suppression of the primary event that unmasks a latent hazard for competing events. In either case, however, intensifying treatment by adding B should not be correlated with a reduced incidence of competing events, and the counterintuitive presence of such a phenomenon is an important indicator of residual bias (given the assumption). This phenomenon has been observed even in randomized trials,^14^ owing to random imbalance of a favorable health characteristics that advantage the A+B arm (particularly smaller trials that are not stratified on such characteristics). More commonly, in non-randomized data, this finding, when it persists after controlling for measurable confounders, is strong evidence of residual confounding, and should be taken into account when making inferences from such data sources.

There are several limitations of this study. Given the above discussion, it is important to note that the approach we describe would not be appropriate when a directional effect on competing events cannot be assumed, such as for many A vs. B study designs. However, the BRACE method can also be applicable to A vs. B designs when the presumption that A does not reduce competing events relative to B is valid (such as when A represents a more invasive treatment approach compared to B). Notably, the proportional relative hazards model treats several key quantities as independent, which is a strong condition that is not always verifiable. Nonetheless, the practical options are limited for investigators who have exhausted standard modeling approaches and still identify a residual confounding problem. While we did not observe critical differences in the performance of the correction method under varying parameters, due to practical limitations we could not examine all permutations or a host of other variables that could theoretically impact our findings. We did note that when the assumption of residual confounding was not valid, the BRACE method resulted in increased potential for type II error resulting from bias toward the null, emphasizing the importance of this assumption with respect to resulting inferences. We also did not take on complex model specifications, such as multiple measured confounders, multi-arm comparisons, dependence between events, or Bayesian approaches. Future work could examine these questions. Many data sources unfortunately lack sufficient information on competing health events, which would preclude applying the BRACE method. However, given the potential for residual confounding in observational data sources, this should be viewed as a limitation of data sources that lack this level of specificity, rather than the method we propose.

## Conclusion

Here we present a novel method (BRACE) to mitigate bias due to residual confounding, based on a proportional relative hazards model. As applied to simulated data, this approach compares favorably to standard methods in terms of lowering bias and model error. Appropriate application of the BRACE method in observational studies on non-randomized data would likely improve effect estimation and inferences.

## Data Availability

Simulated data are able to be generated from the code provided or can be made available upon request to the authors.

---

**Supplementary Table 1.** Additional summary statistics comparing estimation methods under varying scenarios.

| Input Parameters |  |  |  |  |  | Output* |  |  |  |  |  |  |  |  |  |  |  |  |
| --- | --- | --- | --- | --- | --- | --- | --- | --- | --- | --- | --- | --- | --- | --- | --- | --- | --- | --- |
| $\gamma$ | $\kappa^{**}$ | $\omega_0$ | $\Theta_1$ | $\Theta_2$ | $\Theta$ | $\hat{\Theta}_2$ | $\hat{\Theta}$ | | | SE (x 10<sup>-2</sup>; log scale) | | | Bias (x 10<sup>-2</sup>; log scale) | | | MSE (x 10<sup>-2</sup>; log scale) | | |
|  |  |  |  |  |  |  | Standard | BRACE | Hypothetical | Standard | BRACE | Hypothetical | Standard | BRACE | Hypothetical | Standard | BRACE | Hypothetical |
| 0.667 | 0 | 0.50 | 1 | 1 | 1 | 1.002 | 1.005 | 1.007 | 1.005 | 0.458 | 0.452 | 0.456 | 0.492 | 0.726 | 0.494 | 0.456 | 0.411 | 0.462 |
| 0.667 | 0 | 0.50 | 0.5 | 1 | 0.75 | 1.001 | 0.752 | 0.772 | 0.752 | 0.466 | 0.329 | 0.465 | 0.331 | 2.90 | 0.307 | 0.484 | 0.341 | 0.490 |
| 0.667 | 0 | 0.50 | 0.8 | 1.2 | 1 | 1.20 | 1.005 | 1.005 | 1.005 | 0.458 | 0.455 | 0.456 | 0.516 | 0.548 | 0.414 | 0.469 | 0.459 | 0.475 |
| 0.667 | 8 | 0.6 | 1 | 1 | 1 | 0.613 | 0.832 | 0.979 | 1.003 | 0.416 | 0.314 | 0.706 | -18.4 | -2.12 | 0.317 | 3.80 | 0.321 | 0.726 |
| 0.667 | 8 | 0.6 | 0.5 | 1 | 0.7 | 0.612 | 0.550 | 0.727 | 0.661 | 0.445 | 0.261 | 0.715 | -24.1 | 3.76 | -5.76 | 6.25 | 0.373 | 1.05 |
| 0.667 | 8 | 0.6 | 0.84 | 1.25 | 1 | 0.751 | 0.800 | 0.911 | 0.971 | 0.418 | 0.241 | 0.707 | -22.4 | -9.41 | -3.04 | 5.41 | 1.08 | 0.811 |
| 1 | 4 | 0.6 | 0.5 | 1 | 0.7 | 0.709 | 0.587 | 0.714 | 0.666 | 0.433 | 0.215 | 0.521 | -17.6 | 1.93 | -4.97 | 3.51 | 0.257 | 0.742 |
| 1.5 | 4 | 0.6 | 0.5 | 1 | 0.7 | 0.709 | 0.587 | 0.714 | 0.666 | 0.433 | 0.217 | 0.521 | -17.6 | 1.95 | -4.97 | 3.51 | 0.261 | 0.742 |
| 0.667 | 4 | 0.8 | 0.5 | 1 | 0.6 | 0.710 | 0.547 | 0.612 | 0.583 | 0.440 | 0.148 | 0.534 | -9.28 | 2.03 | -2.85 | 1.31 | 0.366 | 0.616 |
| 0.667 | 4 | 0.6 | 0.5*** | 1 | 0.7 | 0.723 | 0.584 | 0.708 | 0.661 | 2.64 | 1.59 | 3.13 | -18.2 | 1.20 | -5.71 | 5.93 | 1.27 | 3.71 |
| *Outputs are expected values across 500 trials per simulation. $\Theta_1$: true effect on event 1. $\Theta_2$: true effect on event 2. $\Theta$: true effect on composite event. $\gamma$: Weibull parameter. $\kappa$: bias parameter. $\omega_0$: baseline relative hazard for event 1 vs. composite event. $\hat{\Theta}_2$: estimated effect on event 2. $\hat{\Theta}$: estimated effect on composite event.<br>**For the bias parameter $\kappa$, when $\kappa = 0$, there is no bias and the baseline hazard for event 2 is unchanged by the biasing factor. For all other values of $\kappa$, when factor is present, the baseline hazard for event 2 is decreased by 1/3 and the log odds of being assigned to the treatment group is multiplied by $\kappa$.<br>***Sample size (n) is 1500 for each trial, except for the bottom row, which had n=250 per trial.<br>BRACE: Bias Reduction through Analysis of Competing Events inverse probability of treatment weighting (not observable in clinical data); SE: standard error; MSE: mean squared error<br>Key findings are presented in highlighted cells. |  |  |  |  |  |  |  |  |  |  |  |  |  |  |  |  |  |  |

## SUPPLEMENTARY INFORMATION

### SAS Macro Used to Generate Simulated Data

~~~
**%MACRO** compsim (D_OUT,nsims,nobs,f,g,b,w1,w2,omegaplus,theta1,theta2,k3);
data gendat;
  do r = **1** to &nsims;
       do j = **1** to &nobs;
                    stream = r*&nsims. + j*&nobs.;
                    call streaminit(stream);
                    *factor is Bernoulli random variable;
                    factor = rand(‘Bernoulli’,&f);
                    *group assignment depends on value of factor and g and b parameters;
                    group = rand(‘Bernoulli’,**1**/(**1**+exp(−(log(&g/(**1**-&g))+log(&b)*factor))));
                    *omega is proportion of baseline composite event hazard attributable to event 1 (lambda01/(lambda01+lambda02);
                    *omegaplus is ratio of baseline event hazards lambda01/lambda02 (fixed effect);
                    *Weibull values depend on shape (a) and scale (b) parameters;
                    *f(t) = (a/(b^a))*(t^(a-1))*(exp(−(t/b)^a));
                    *h(t) = (b^(−a))*a*(t^(a-1));
                    *a < 1 ==> hazard is decreasing;
                    *proportional effects on h(t) will multiply scaling parameter b^-a (alternate Weibull parametrization);
                    *theta1 and omegaplus are multipliers on the baseline hazard for event 1;
                    *theta2 and k3 are multipliers on the baseline hazard for event 2;
                    *for a multiplier M, the scaling factor S = (1/M)^(1/w) = M^(−1/w);
                    *default scale = 1;
                    scale1 = (&omegaplus*(**1**-group*(**1**-&theta1)))**(−**1**/&w1);
                    scale2 = ((**1**-group*(**1**-&theta2))*(**1**-factor*(**1**-&k3)))**(−**1**/&w2);
                    *scale1 = 1;
                    *scale2 = 1;
                    *if group = 1 then do;
                    * scale1 = (1/(&theta1))**(1/&w1);
                    * scale2 = (1/(&theta2))**(1/&w2);
                    * end;
                    *if factor = 1 then scale2 = (1/(&k3))**(1/&w2);
                    t1 = rand(‘WEIBull’, &w1, scale1);
                    t2 = rand(‘WEIBull’, &w2, scale2);
                    time = min(t1, t2);
                    if (factor=**0** & group=**0** & (t1 < t2)) then report=**1**;
                    else if (factor=**0** & group=**0** & (t1 > t2)) then report=**2**;
                    else report=**0**;
                    event1 = **0**;
                    event2 = **0**;
                    if t1 < t2 then event1=**1**;
                    else event2=**1**;
                    * impose random censoring;
                               censored = (ranuni(**834332353**)>.**66**);
                    if censored then do;
                                        event1=**0**;
                                        event2=**0**;
                                        end;
                            output;
                    end;
               end;
proc sort data = gendat;
       by group;
       run;
data temp;
       set gendat;
       drop j;
event=**0**;
if event1=**1** then event=**1**;
if event2=**1** then event=**1**;
pfs_ci=**0**;
if event1=**1** then pfs_ci=**1**;
if event2=**1** then pfs_ci=**2**;
*scale and map event times;
time = round(**6000**\*time)/**100** + **6**;
trueomega = &omegaplus./(&omegaplus. + **1** - &f. + &f. * &k3.);
truetheta = (&theta1.*&omegaplus. + &theta2. - &theta2. * &f. + &theta2. * &f. * &k3.)/(&omegaplus. + **1** - &f. + &f. * &k3.);
run;
data &D_OUT;
               set temp;
               run;
PROC EXPORT DATA= work.&D_OUT.
            OUTFILE= “*”
            DBMS=TAB LABEL REPLACE;
     PUTNAMES=YES;
RUN;
**%mend** compsim;
~~~

### R Code for Analysis of Simulated Data

~~~
library(cmprsk)
library(gcerisk)
library(psych)
library(plyr)
library(boot)
library(rsimsum)
library(ggplot2)
library(Hmisc)
library(dplyr)
library(purrr)
library(stringr)
require(MASS)
require(WeightIt)
###########################################################################
#
# DEFINE FUNCTIONS FOR BOOTSTRAP ROUTINES
#
###########################################################################
#function for bootstrap estimates
superfunct<- function(d,index){
   d2<-d[index,]
   dtemp0=subset(d2,group==0)
   dttx <- basehaz(coxph(Surv(dtemp0$time,dtemp0$event) ∼ 1))[,2]
   dind <- which(abs(log(dttx) - mean(log(dtemp0$time))) == min(abs(log(dttx)-mean(log(dtemp0$time))), na.rm=TRUE))
   ddwf <- 1-mean(basehaz(coxph(Surv(dtemp0$time,dtemp0$event1)∼1))[dind,1])/mean(basehaz(coxph(Surv(dtemp0$time,dtemp0$event)∼1))[dind,1])
   dtheta <- summary(coxph(Surv(d2$time,d2$event) ∼ d2$group))$coef[2]
   dtheta2 <- summary(coxph(Surv(d2$time,d2$event2) ∼ d2$group))$coef[2]
   depsilon = ddwf*(1-dtheta2)
   dthetaM <- as.numeric(dtheta)+ depsilon
   ithetaM <- exp(coxph(Surv(time, event) ∼ group + factor, data=d2, weights=weightit(group ∼ factor, data=d2, method=“ps”, estimand=“ATT”, stabilize=TRUE)$weights)$coef[1])
   #return(dthetaM)
   #return(dtheta)
   return(ithetaM)
}
#function for Studentized bootstrap estimates
get_r_var <- function(grv_d, grv_index, its) {
   grv_d2 <- grv_d[grv_index,]
   grv_temp0 = subset(grv_d2, grv_d2$group==0)
   grv_ttx <- basehaz(coxph(Surv(grv_temp0$time,grv_temp0$event) ∼ 1))[,2]
   grv_ind <- which(abs(log(grv_ttx) - mean(log(grv_temp0$time))) == min(abs(log(grv_ttx)-
mean(log(grv_temp0$time))), na.rm=TRUE))
   grv_dwf <- 1-
mean(basehaz(coxph(Surv(grv_temp0$time,grv_temp0$event1)∼1))[ind,1])/mean(basehaz(coxph(Surv(grv_temp0$t
ime,grv_temp0$event)∼1))[ind,1])
   grv_theta <- summary(coxph(Surv(grv_d2$time,grv_d2$event) ∼ grv_d2$group))$coef[2]
   grv_theta2 <- summary(coxph(Surv(grv_d2$time,grv_d2$event2) ∼ grv_d2$group))$coef[2]
   log.boot.thetaM <- log(as.numeric(grv_theta)+ grv_dwf*(1-grv_theta2))
   grv_n = nrow(grv_d)
   grv_v <- boot(grv_d, R = its, statistic = superfunct) %>% pluck(“t”) %>% var(na.rm = TRUE)
c(log.boot.thetaM, grv_v)
}
#function for Monte Carlo estimates
montecarlo <- function(d){
   temp0=subset(d,d$group==0)
   ttx <- basehaz(coxph(Surv(temp0$time,temp0$event) ∼ 1))[,2]
   ind <- which(abs(log(ttx) - mean(log(temp0$time))) == min(abs(log(ttx)-mean(log(temp0$time))), na.rm=TRUE))
   theta <- summary(coxph(Surv(d$time,d$event) ∼ d$group))$coef[2]
   thetase <- summary(coxph(Surv(d$time,d$event) ∼ d$group))$coef[3]
   theta2 <- summary(coxph(Surv(d$time,d$event2) ∼ d$group))$coef[2]
   theta2se <- summary(coxph(Surv(d$time,d$event2) ∼ d$group))$coef[3]
   ev1 <- (survfit(coxph(Surv(temp0$time,temp0$event1)∼1), ctype = 1))
   ev <- (survfit(coxph(Surv(temp0$time,temp0$event)∼1), ctype = 1))
   ev1.haz <- ev1$cumhaz[ind]
   ev.haz <- ev$cumhaz[ind]
   ev1.se <- ev1$std.err[ind]
   ev.se <- ev$std.err[ind]
   B <- array(0, c(studreps))
   for(b in 1:studreps){
      set.seed(b*max(d$r))
      B[b] = theta + (1-rnorm(1,mean=ev1.haz,sd=ev1.se)/rnorm(1,mean=ev.haz,sd=ev.se))*(1-rnorm(1,mean=theta2,sd=theta2se))
      }
      return(B)
}
###########################################################################
#
# PROCESS SIMULATED DATA
#
# G: FOR EACH SIM, STORES VECTOR OF TRIAL-LEVEL STATS
#
# H: FOR EACH SIM, STORES 3 THETA/SE ESTIMATES - BIASED, METHOD-ADJ, REG-ADJ
#
# J: FOR EACH SIM, STORES VECTOR OF MEAN VALUES ACROSS ALL TRIALS
#
###########################################################################
startsim=32
endsim=33
nsims=endsim-startsim+1
trials = 500
bootreps = 500
studreps = 1000
#initialize output matrices
  #G has 1 record per trial
  G <- array(0, c(trials,43))
  colnames(G)=c(“r”,”theta”,”theta1”,”theta2”,”%factor”,”%group”,”%factor_G0”,”%factor_G1”,”%group_F0”,
             “%group_F1”,”omega”,”omega+”,”epsilon”,”thetaB_95L”,”thetaB”,”thetaB_95U”,”thetaM”,
             “thetaR_95L”,”thetaR”,”thetaR_95U”,”thetaS0_95L”,”thetaS0”,”thetaS0_95U”,”thetaS1_95L”,
“thetaS1”,”thetaS1_95U”,”diffM_R”,”truetheta”,”ivar1”,”ivar2”,”thetaI”,”trueomega”,
             “p_unadj”,”r_unadj”, “diffM_T”,”p_adj”,”r_adj”,”thetaM_95L”,”thetaM_95U”, “E.var.thetaM”,
             “p_method”,”r_method”,”log_boot_thetaM”)
#H has 3 records per trial per sim
H <- array(0, c(5*trials, 7, nsims))
colnames(H)=c(“dataset”,”n”,”baseline”,”theta”,”se”,”model”,”epsilon”)
#J has vector of mean values, 1 record per sim
J <- as.data.frame(array(0, c(44,nsims)))
rownames(J)=c(“theta”,”theta1”,”theta2”,”%factor”,”%group”,”%factor_G0”,”%factor_G1”,”%group_F0”,
             “%group_F1”,”omega”,”omega+”,”epsilon”,”thetaB_95L”,”thetaB”,”thetaB_95U”,”thetaM”,
             “thetaR_95L”,”thetaR”,”thetaR_95U”,”thetaS0_95L”,”thetaS0”,”thetaS0_95U”,”thetaS1_95L”,
             “thetaS1”,”thetaS1_95U”,”diffM_R”,”truetheta”,”ivar1”,”ivar2”,”thetaI”,”trueomega”,
             “p_unadj”,”r_unadj”, “diffM_T”,”p_adj”,”r_adj”, “thetaM_95L”,”thetaM_95U”,
             “E.var.thetaM”, “p_method”,”r_method”,”log_boot_thetaM”,”var.thetaM”,”mse.thetaM”)
#start loop
  for (z in 1:nsims) {
data = simlist[[startsim+z-1]]
#assign(“data”,get(paste0(‘sim’,startsim+z-1)))
subset(data, r <= trials) -> data
H[,2,z] = nrow(data)/trials
H[,3,z] = max(data$base)
for (i in 1:max(data$r)) {
     #Matrix Labels
     H[(5*(i-1)+1),1,z] = i
     H[(5*(i-1)+2),1,z] = i
     H[(5*(i-1)+3),1,z] = i
     H[(5*(i-1)+4),1,z] = i
     H[(5*(i-1)+5),1,z] = i
     G[i,1] = i
     H[(5*(i-1)+1),6,z] = c(“Unadjusted”)
     H[(5*(i-1)+2),6,z] = c(“Method”)
     H[(5*(i-1)+3),6,z] = c(“IPTW”)
     H[(5*(i-1)+4),6,z] = c(“Strat0”)
     H[(5*(i-1)+5),6,z] = c(“Strat1”)
     #Working Data Sets
     temp = subset(data,r==i)
     temp0 = subset(temp,group==0)
     temp2=subset(temp,factor==0)
     temp3=subset(temp,factor==1)
     #Mean Values for Each Sample Trial
     G[i,5] <- mean(temp$factor)
     G[i,6] <- mean(temp$group)
     G[i,7] <- mean(subset(temp, group==0)$factor)
     G[i,8] <- mean(subset(temp, group==1)$factor)
     G[i,9] <- mean(subset(temp, factor==0)$group)
     G[i,10] <- mean(subset(temp, factor==1)$group)
     #Unadjusted Effects on Event, Event1, Event2
     G[i,2] <- summary(coxph(Surv(temp$time,temp$event) ∼ temp$group))$coef[2]
     G[i,3] <- summary(coxph(Surv(temp$time,temp$event1) ∼ temp$group))$coef[2]
     G[i,4] <- summary(coxph(Surv(temp$time,temp$event2) ∼ temp$group))$coef[2]
     #Stratified Estimates
     G[i,21] <- summary(coxph(Surv(temp2$time,temp2$event) ∼ temp2$group))$conf[1,3]
     G[i,22] <- summary(coxph(Surv(temp2$time,temp2$event) ∼ temp2$group))$coef[1,2]
     G[i,23] <- summary(coxph(Surv(temp2$time,temp2$event) ∼ temp2$group))$conf[1,4]
     H[(5*(i-1)+4),4,z] = G[i,22]
     H[(5*(i-1)+4),5,z] = summary(coxph(Surv(temp2$time,temp2$event) ∼ temp2$group))$coef[1,3]
     G[i,24] <- summary(coxph(Surv(temp3$time,temp3$event) ∼ temp3$group))$conf[1,3]
     G[i,25] <- summary(coxph(Surv(temp3$time,temp3$event) ∼ temp3$group))$coef[1,2]
     G[i,26] <- summary(coxph(Surv(temp3$time,temp3$event) ∼ temp3$group))$conf[1,4]
     H[(5*(i-1)+5),4,z] = G[i,25]
     H[(5*(i-1)+5),5,z] = summary(coxph(Surv(temp3$time,temp3$event) ∼ temp3$group))$coef[1,3]
     #Unadjusted Estimates
     G[i,14] <- summary(coxph(Surv(temp$time,temp$event) ∼ temp$group))$conf[1,3]
     G[i,15] <- summary(coxph(Surv(temp$time,temp$event) ∼ temp$group))$coef[1,2]
     G[i,16] <- summary(coxph(Surv(temp$time,temp$event) ∼ temp$group))$conf[1,4]
     H[(5*(i-1)+1),4,z] = G[i,15]
     H[(5*(i-1)+1),5,z] = summary(coxph(Surv(temp$time,temp$event) ∼ temp$group))$coef[1,3]
     #IPTW (Hypothetical) Estimates
     G[i,18] <- summary(coxph(Surv(time, event) ∼ group + factor, data=temp, weights=weightit(group ∼ factor, data=temp, method=“ps”, estimand=“ATT”, stabilize=TRUE)$weights))$conf[1,3]
     G[i,19] <- exp(coxph(Surv(time, event) ∼ group + factor, data=temp, weights=weightit(group ∼ factor, data=temp, method=“ps”, estimand=“ATT”, stabilize=TRUE)$weights)$coef[1])
     G[i,20] <- summary(coxph(Surv(time, event) ∼ group + factor, data=temp, weights=weightit(group ∼ factor, data=temp, method=“ps”, estimand=“ATT”, stabilize=TRUE)$weights))$conf[1,4]
     H[(5*(i-1)+3),4,z] = G[i,19]
     H[(5*(i-1)+3),5,z] = summary(coxph(Surv(time, event) ∼ group + factor, data=temp,
     weights=weightit(group ∼ factor, data=temp, method=“ps”, estimand=“ATT”,
     stabilize=TRUE)$weights))$coef[1,4]
     #BRACE Adjusted Estimates
     ttx <- basehaz(coxph(Surv(temp0$time,temp0$event) ∼ 1))[,2]
     ind <- which(abs(log(ttx) - mean(log(temp0$time))) == min(abs(log(ttx)-mean(log(temp0$time))), na.rm=TRUE))
     G[i,11] <-
basehaz(coxph(Surv(temp0$time,temp0$event1)∼1))[ind,1]/basehaz(coxph(Surv(temp0$time,temp0$event)∼1))[ind,1]
     G[i,12] <-
basehaz(coxph(Surv(temp0$time,temp0$event1)∼1))[ind,1]/basehaz(coxph(Surv(temp0$time,temp0$event2)∼1))[ind,1]
     G[i,13] <- (1-G[i,11])*(1-G[i,4])
     G[i,17] = ifelse((G[i,13] > 0) & (G[i,16] < 1),mean(montecarlo(temp)),G[i,15])
     H[(5*(i-1)+2),4,z] = G[i,17]
     H[(5*(i-1)+2),5,z] = ifelse((G[i,13] > 0) & (G[i,16] < 1),sd(montecarlo(temp)),H[(5*(i-1)+1),5,z])
     H[(5*(i-1)+1),7,z] = G[i,17] - G[i,15]
     H[(5*(i-1)+2),7,z] = G[i,17] - G[i,15]
     H[(5*(i-1)+3),7,z] = G[i,17] - G[i,15]
     H[(5*(i-1)+4),7,z] = G[i,17] - G[i,15]
     H[(5*(i-1)+5),7,z] = G[i,17] - G[i,15]
     G[i,40] <- var(montecarlo(temp))
     G[i,38] <- G[i,17] - 1.96*sd(montecarlo(temp))
     G[i,39] <- G[i,17] + 1.96*sd(montecarlo(temp))
     G[i,43] <- log(mean(montecarlo(temp)))
     #Method vs. IPTW
     G[i,27] <- G[i,17] - G[i,19]
     #Is true theta in Sim Interval?
     G[i,28] <- mean(temp$truetheta)
     G[i,33] <- (G[i,14] <= G[i,28]) & (G[i,16] >= G[i,28])+0
     G[i,36] <- (G[i,18] <= G[i,28]) & (G[i,20] >= G[i,28])+0
     G[i,41] <- (G[i,38] <= G[i,28]) & (G[i,39] >= G[i,28])+0
     #Method vs. True Theta
     G[i,35] <- G[i,17] - G[i,28]
     #Is Null in Sim Interval?
     G[i,32] <- mean(temp$trueomega)
     G[i,34] <- (G[i,14] > 1) | (G[i,16] < 1)+0
     G[i,37] <- (G[i,18] > 1) | (G[i,20] < 1)+0
     G[i,42] <- (G[i,38] > 1) | (G[i,39] < 1)+0
     #Inverse variance weighted estimates
     G[i,29] <- 1/((summary(coxph(Surv(temp2$time,temp2$event) ∼ temp2$group))$coef[3])^2)
     G[i,30] <- 1/((summary(coxph(Surv(temp3$time,temp3$event) ∼ temp3$group))$coef[3])^2)
     G[i,31] <- (G[i,29]*G[i,22]+G[i,30]*G[i,25])/(G[i,29]+G[i,30])
#close 2nd for loop
}
as.data.frame(G) -> G
assign(paste0(“G”, startsim+z-1),G)
for (i in 2:ncol(G)) {
J[i-1,z] <- mean(G[,i])
}
J[ncol(G),z] = var(G[,17])
J[ncol(G)+1,z] = mean((G[,17]-G[,28])^2)
assign(paste0(“H”, startsim+z-1),as.data.frame(H[,,z]))
assign(paste0(“J”, startsim+z-1),as.data.frame(J[,z]))
write.table(assign(paste0(“J”, startsim+z-1),J[,z]), paste0(paste0(“*”, startsim+z-1),”.txt”), sep=“\t”)
write.table(assign(paste0(“G”, startsim+z-1),G), paste0(paste0(“*”, startsim+z-1),”.txt”), sep=“\t”)
write.table(assign(paste0(“H”, startsim+z-1),H[,,z]), paste0(paste0(“*”, startsim+z-1),”.txt”), sep=“\t”)#close 1st for loop
}
##################################################
#
# GENERATE SIM PLOTS
#
##################################################
#call sim number here
sim <- 33
#read in Ordered sim output
Glist <- list()
Hlist <- list()
Jlist <- list()
for (j in sim:sim) {
    assign(paste0(“G”,j), read.table(paste0(paste0(“*”, j),”.txt”),header=TRUE,sep=“\t”))
    Glist[[j]] <- get(paste0(“G”,j))
assign(paste0(“H”,j), read.table(paste0(paste0(“*”, j),”.txt”),header=TRUE,sep=“\t”))
    Hlist[[j]] <- get(paste0(“H”,j))
    assign(paste0(“J”,j), read.table(paste0(paste0(“*”, j),”.txt”),header=TRUE,sep=“\t”))
Jlist[[j]] <- get(paste0(“J”,j))
rownames(Jlist[[j]]) =
c(“theta”,”theta1”,”theta2”,”%factor”,”%group”,”%factor_G0”,”%factor_G1”,”%group_F0”,”%group_F1”,”omega”,”omega+”,”epsilon”,”thetaB_95L”,”thetaB”,”thetaB_95U”,”thetaM”,”thetaR_95L”,”thetaR”,”thetaR_95U”,”thetaS0_95L”,”thetaS0”,”thetaS0_95U”,”thetaS1_95L”,”thetaS1”,”thetaS1_95U”,”diffM_R”,”truetheta”,”ivar1”,”ivar2”,”thetaI”,”trueomega”,”p_unadj”,”r_unadj”, “diffM_T”,”p_adj”,”r_adj”, “thetaM_95L”,”thetaM_95U”, “E.var.thetaM”,”p_method”,”r_method”,”log_boot_thetaM”,”var.thetaM”,”mse.thetaM”)
colnames(Glist[[j]])=c(“r”,”theta”,”theta1”,”theta2”,”%factor”,”%group”,”%factor_G0”,”%factor_G1”,”%group_F0”,”%group_F1”,”omega”,”omega+”,”epsilon”,”thetaB_95L”,”thetaB”,”thetaB_95U”,”thetaM”,”thetaR_95L”,”thetaR”,”thetaR_95U”,”thetaS0_95L”,”thetaS0”,”thetaS0_95U”,”thetaS1_95L”,”thetaS1”,”thetaS1_95U”,”diffM_R”,”truetheta”,”ivar1”,”ivar2”,”thetaI”,”trueomega”,”p_unadj”,”r_unadj”,”diffM_T”,”p_adj”,”r_adj”,”thetaM_95L”,”thetaM_95U”, “E.var.thetaM”,”p_method”,”r_method”,”log_boot_thetaM”)
}
J0 <- as.data.frame(Jlist[[sim]])
G0 <- as.data.frame(Glist[[sim]])
H0 <- as.data.frame(Hlist[[sim]])
H0$theta = log(as.numeric(H0$theta))
H0$epsilon = as.numeric(H0$epsilon)
H0$se = as.numeric(H0$se)
colnames(H0)=c(“dataset”,”n”,”baseline”,”log(theta)”,”se”,”model”,”epsilon”)
H0$model[H0$model == “Method”] <- “BRACE”
H0$model[H0$model == “Unadjusted”] <- “Standard”
H0$model[H0$model == “IPTW”] <- “Hypothetical”
H0$baseline = simlist[[sim]]$base[1]
Htrue=log(Jlist[[sim]][27,])
Htrue
H0 = subset(H0,model %in% c(“Standard”,”BRACE”,”Hypothetical”))
for (i in 2:ncol(G0)) {
J0[i-1,1] <- mean(G0[,i])
}
J0[ncol(G0),1] = var(G0[,17])
J0[ncol(G0)+1,1] = mean((G0[,17]-G0[,28])^2)
s1 <- simsum(
    data = H0, estvarname = “log(theta)”, se = “se”, true = Htrue,
    methodvar = “model”, ref=“IPTW”, by = c(“n”, “baseline”), x = TRUE
)
#Figure 1
autoplot(s1, type = “est_ridge”) + theme(axis.text.y=element_blank()) + viridis::scale_fill_viridis(discrete = TRUE) + viridis::scale_colour_viridis(discrete = TRUE)
#Figure 2
h0=as.data.frame(cbind(H0$model,H0$‘log(theta)’))
as.numeric(h0$V2) - log(Jlist[[sim]][27,]) -> h0$V2
as.factor(h0$V1) -> h0$V1
fill <- “#4271AE”
lines <- “#1F3552”
p10 <- ggplot(h0, aes(x = V1,y = V2)) +
    geom_boxplot(colour = lines, fill = fill,
size = 1) +
    scale_y_continuous(name = “Bias”,
breaks = seq(−0.4, 0.3, 0.1),
limits=c(−0.4, 0.3)) +
    scale_x_discrete(name = “Model”) +
ggtitle(““) +
theme_bw() +
theme(panel.grid.major = element_line(colour = “#d3d3d3”),
      panel.grid.minor = element_blank(),
      panel.border = element_blank(),
      panel.background = element_blank(),
      plot.title = element_text(size = 14, family = “Tahoma”, face = “bold”),
      text=element_text(family = “Tahoma”),
      axis.title = element_text(face=“bold”),
      axis.text.x = element_text(colour=“black”, size = 11),
      axis.text.y = element_text(colour=“black”, size = 9),
      axis.line = element_line(size=0.5, colour = “black”))
p10
#autoplot(summary(s1), type = “forest”, stats = “bias”)
#Figure 3
p11 <- autoplot(summary(s1), type = “forest”, stats = “mse”, colour = lines, fill = fill, size = 1) +
    scale_y_continuous(name = “MSE”,
                       breaks = seq(0, .040, 0.005),
                       limits=c(0, .04)) +
    scale_x_discrete(name = “Model”) +
    ggtitle(““) +
    theme_bw() +
    theme(panel.grid.major = element_line(colour = “#d3d3d3”),
          panel.grid.minor = element_blank(),
          panel.border = element_blank(),
          panel.background = element_blank(),
          plot.title = element_text(size = 14, family = “Tahoma”, face = “bold”),
          text=element_text(family = “Tahoma”),
          axis.title = element_text(face=“bold”),
          axis.text.x = element_text(colour=“black”, size = 11),
          axis.text.y = element_text(colour=“black”, size = 9),
          axis.line = element_line(size=0.5, colour = “black”))
p11
#Figure 4
autoplot(s1, type = “zip”)
~~~

